# Autoantibodies neutralizing type I interferons underlie a third of cases of Chikungunya virus encephalitis or myelitis

**DOI:** 10.64898/2026.07.23.26358609

**Authors:** Vu L. Tran, Adrian Gervais, Marie-Mechtilde Champeaux, Maria Paula de Souza Sampaio, Sylvie Abel, Isabelle Calmont, Mateus Santana do Rosario, Laire Schidlowski, Jordan D. Simione, Pedro Augusto Alves, Anne Puel, Paul Bastard, Laurent Abel, Carolina Prando, Rafael Freitas de Oliveira Franca, Isadora Cristina de Siqueira, André Cabié, Aurélie Cobat, Shen-Ying Zhang, Jean-Laurent Casanova

**Author notes:** equal contribution (Vu L. Tran, Adrian Gervais). equal contribution (Rafael Freitas de Oliveira Franca, Isadora Cristina de Siqueira, André Cabié). equal contribution (Aurélie Cobat, Shen-Ying Zhang, Jean-Laurent Casanova).

## Abstract

Chikungunya virus (CHIKV) infection is typically not life-threatening but may, in rare cases, affect the central nervous system (CNS). In three cohorts of confirmed CHIKV cases from Martinique (French overseas territory) and Brazil (patients aged 0–89 years, *n* = 245), 20 patients had CNS infection (aged 17–87 years). Autoantibodies neutralizing type I interferons (AAN-I-IFN) were found in 35% of patients (4 male and 3 female, aged 17–87 years) with encephalitis (4/15), encephalomyelitis (1/2), or myelitis (2/3), but were absent in 225 patients without CNS infection. They neutralized high concentrations (10–1,000 ng/mL) of IFN-α2, IFN-α8, and IFN-ω, and the antibodies of one patient also neutralized IFN-β. All samples also neutralized the other 10 IFN-α subtypes (at least 100 pg/mL). This combination of blood autoantibodies occurs in ∼0.02% and 0.6% of healthy individuals under and over 70 years of age, respectively. The presence of AAN-I-IFN before CHIKV infection therefore increased the risk of CNS disease ∼850-fold relative to the general population.

## Introduction

Chikungunya virus (CHIKV) is an *Alphavirus* from the *Togoviridae* family, transmitted to humans primarily by *Aedes aegypti* and *Aedes albopictus* mosquitoes (1). Until the early 2000’s, CHIKV caused outbreaks only in Africa and Asia, but its geographic range has since expanded, with notable outbreaks in the Americas, Oceania, and Europe (2, 3). Chikungunya is, therefore, becoming a significant burden on healthcare systems worldwide. CHIKV can cause outbreaks in areas in which no prior immunity has been established, as in the Americas in 2013 (2) and China in 2025 (4). The first locally transmitted cases of CHIKV infection were reported in the USA in 2014 (2), with 3.7 million suspected or confirmed infections occurring in the Americas over the decade that followed (5). Infection rates can be very high, with reports that 10-70% of individuals in endemic regions may be infected (6). One recent study estimated the annual number of human infections globally at about 35 million, with most cases occurring in Southeast Asia, Africa, and the Americas (7).

Human CHIKV infection principally causes high fever with polyarthralgia (8). Severe infections are rare, with a fatality rate of ∼0.3% in the infected general population (8). In severe cases, CHIKV can lead to life-threatening central nervous system (CNS) infection (encephalitis, meningoencephalitis, myelitis), and/or severe post-viral complications of the peripheral nervous system (Guillain-Barré Syndrome) (9). The risk of severe acute CHIKV infection requiring hospitalization increases substantially with age and differs between the sexes: the risk of developing severe CHIKV infection in individuals over the age of 60 years is about 20 times higher than that in individuals under the age of 60 years and about 80 times higher than that in individuals aged 20–29 years, and the odds of death from CHIKV infection in male patients are twice that in female patients (10). Interestingly, the incidence of CHIKV encephalitis follows a U-shaped distribution with two peaks, one in infancy (<1 year of age) and the other in the elderly (>65 years of age) (11, 12). The mechanisms underlying severe neurological CHIKV in humans remain unclear.

We recently reported that autoantibodies (auto-Abs) neutralizing type I interferons (IFN-I) (AAN-I-IFN) are common and strong determinants of a growing number of life-threatening viral infections (13–19). During the 2025 vaccination campaign in response to the recent CHIKV outbreak on Reunion Island, five unrelated elderly individuals developed severe adverse reactions to the live attenuated vaccine VLA1553 (20). Three had encephalitis and all three had high levels of AAN-I-IFN in blood and cerebral spinal fluid (CSF); the other two had a severely impaired general state without encephalitis and with no detectable AAN-I-IFN. We, thus, hypothesized that AAN-I-IFN might also underlie severe forms of naturally occurring CHIKV infection.

## Results

### Three Cohorts of CHIKV-infected Individuals

We studied 245 individuals with CHIKV infection from Martinique (A French overseas territory, *n* = 204, collected in 2014) and Brazil (two independent cohorts, one from the Northeast, collected in 2014–2016 (21), another from Bahia State in the Northeast, collected from 2020–2025 (22), *n* = 41 in total) (Fig. 1*A*). Ten healthy donors from Brazil were included as controls. The CHIKV-infected patients were classified into two different groups based on their clinical manifestations: mild (*n* = 225) and severe central nervous system (CNS) infection (*n* = 20 in total), including myelitis (*n* = 3), encephalitis (*n* = 15), and encephalomyelitis (*n* = 2). Most patients with mild symptoms were managed at home without hospitalization. All 20 patients with CNS infection were hospitalized in an intensive care unit (ICU), and one man in his 80’s died after 33 days due to neurological deterioration. Plasma/serum samples were collected from all except four patients during the acute phase of infection (median delay between symptom onset and sampling = 4 days (0−31 days)). We also had access to cerebrospinal fluid (CSF) from six of the encephalitis cases and two of the encephalomyelitis cases from Brazil.

**Fig. 1.**
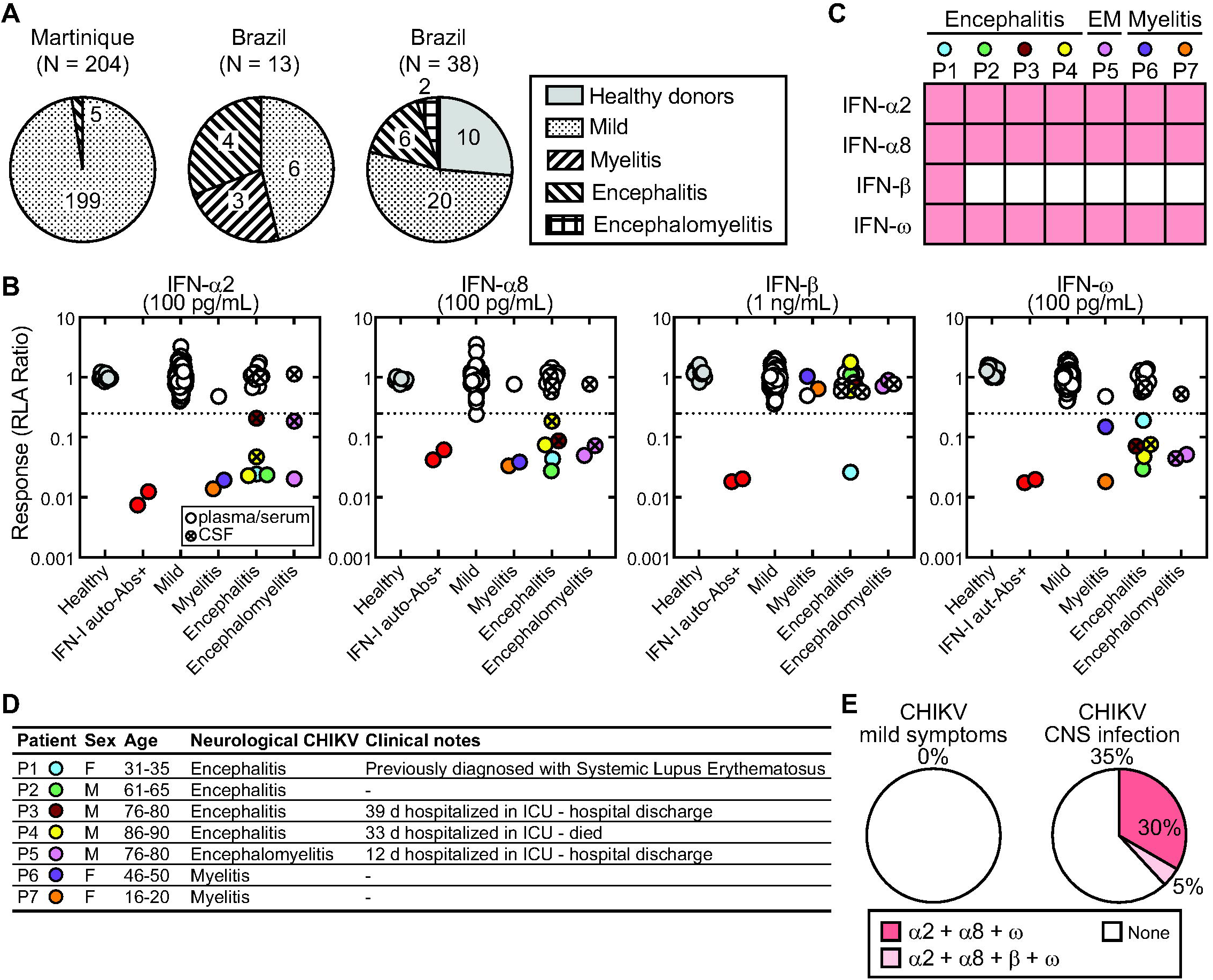
Autoantibodies neutralizing IFN-I in CHIKV-infected patients with severe encephalitis/myelitis. (*A*) 245 CHIKV-infected patients from Martinique (France) and Brazil were classified into different groups on the basis of clinical manifestations. (*B*) Detection of IFN-I autoantibodies (auto-Abs) in a Luciferase-based neutralization assay; RLA: relative luciferase activity. Samples with an RLA ratio < 0.25 were considered neutralizing. (*C*) Heatmap depicting the presence of IFN-I auto-Abs in seven individuals. EM: Encephalomyelitis; red: positive, white: negative. (*D*) General clinical characteristics of seven IFN-I auto-Ab-positive patients. ICU: intensive care unit. (*E*) Proportion of IFN-I neutralized in CHIKV-infected patients with neurological complications versus those with mild symptoms.

### Autoantibodies Neutralizing IFN-I in CHIKV-infected Patients with Severe Neurological Complications

We investigated the presence of AAN-I-IFN in these patients, using a previously described luciferase-based neutralization assay in A549 IFN Reporter (AIR) cells (23). The neutralization activity of the patient samples served as a surrogate for the presence of AAN-I-IFN. Plasma or serum samples were first tested at a 1:20 dilution for their ability to neutralize 100 pg/mL IFN-α2, IFN-α8, or IFN-ω, or 1 ng/mL IFN-β. Strikingly, six of the 16 patients (37.5%) with severe CNS infection carried circulating AAN-I-IFN effectively neutralizing at least three different types of IFN-I (IFN-α2, IFN-α8 and IFN-ω) (Fig. 1 *B*-*D*). We further tested the CSF (1:20 dilution) of eight patients with severe CNS infection, including two with AAN-I-IFN circulating in the blood, two without circulating AAN-I-IFN and four for whom no serum or plasma was available. Three of the eight patients (37.5%) had CSF AAN-I-IFN, including the two with blood AAN-I-IFN and one of unknown blood AAN-I-IFN status. In total, seven patients carried AAN-I-IFN in blood and/or CSF. They were aged from 17 to 87 years; four were male and three were female. One woman (P1) in her 30’s had previously been diagnosed with systemic lupus erythematosus (SLE), a known etiology of AAN-I-IFN (24), but the other patients were previously healthy. Four of these patients had encephalitis (4 of 15), one had encephalomyelitis (1 of 2), and two had myelitis (2 of 3).

P1 was the only patient whose samples neutralized all types of IFN-I tested (IFN-α2, IFN-α8, IFN-β, and IFN-ω). The samples of three other encephalitis patients (P2-P4) neutralized IFN-α2, IFN-α8, and IFN-ω. Only CSF was available from P3, and it effectively blocked the activity of IFN-α2, IFN-α8, and IFN-ω. Interestingly, both the serum and CSF samples of P4 and P5 displayed neutralizing activity, indicating the circulation of AAN-I-IFN in the CNS, neutralizing IFN-I at concentrations at least as high as 2,000 pg/mL *in vivo* (equivalent to 100 pg/mL neutralized by samples diluted 1:20). P3 and P4 were men and in their 80’s; both were hospitalized in the ICU, for 39 and 33 days, respectively. P3 was later discharged, but P4 died. P5 was a man who developed encephalomyelitis and was discharged after 12 days of hospitalization in the ICU. The serum/CSF samples of two female patients with myelitis (P6 and P7) neutralized IFN-α2, IFN-α8, and IFN-ω, but not IFN-β. P6 was in her 40’s and P7 was in 16–20-year-old range. By contrast, none of the 225 cases with mild symptoms had detectable AAN-I-IFN circulating in the blood (Fig. 1*B*). IFN-I auto-Abs were found to underlie a third of severe cases of CHIKV CNS infection (Fig. 1*E*).

### Highly Potent Autoantibodies Neutralizing Supraphysiological Levels of IFN-I

Seven patients with AAN-I-IFN developed CNS complications of various degrees of severity, ranging from prolonged hospitalization to death. We therefore hypothesized that the patients with the most potent and/or abundant AAN-I-IFN might have more unfavorable outcomes. We then tested all positive samples, serially diluted (from 1:200 to 1:200,000), in the presence of 100 pg/mL IFN-α2, IFN-α8, or IFN-ω or 1 ng/mL IFN-β, to determine the highest equivalent concentration of IFN-I neutralized by these samples. The samples of all patients neutralized IFN-α2 and IFN-α8 when diluted 1:200 (or 5×10^−3^) (Fig. 2 *A* and *B*). The samples of P1 neutralized both IFN-α2 and IFN-ω at a 1:2,000 (or 5×10^−4^) dilution and both IFN-α8 and IFN-β at a 1:200 dilution. The samples of P6, a patient with myelitis, neutralized both IFN-α2 and IFN-α8 at a 1:200 dilution but was unable to neutralize IFN-ω at this dilution. By contrast, plasma from P7, who had myelitis, retained its neutralization activity even at a dilution of 1:20,000 (or 5×10^−5^) for IFN-α2 and IFN-α8 or 1:200,000 (or 5×10^−6^) for IFN-ω. Remarkably, the serum of P4 neutralized IFN-α2 and IFN-ω even at a dilution of 1:200,000 and IFN-α8 at a dilution of 1:20,000. This is equivalent to the 1:20 dilution of serum neutralizing 1,000 ng/mL IFN-α2 and IFN-ω and 100 ng/mL IFN-α8. Strikingly, this patient (P4), with the most potently neutralizing auto-Abs of all patients tested, was the only one who died after infection. Interestingly, the CSF of P4 lost its neutralization activity when diluted by a factor of more than 100 times relative to the starter, possibly reflecting the presence of smaller amounts of IgG in the CSF than in plasma. By contrast, both the serum and CSF of P5 retained their neutralization activity when diluted 1:20,000 for IFN-α2 (equivalent of 100 ng/mL IFN-α2 *in vivo*) or 1:2,000 for IFN-α8 and IFN-ω (equivalent of 10 ng/mL IFN-α8/IFN-ω *in vivo*). These data indicate that the potency and/or abundance of circulating AAN-I-IFN is not always lower in the CSF than in the blood.

**Fig. 2.**
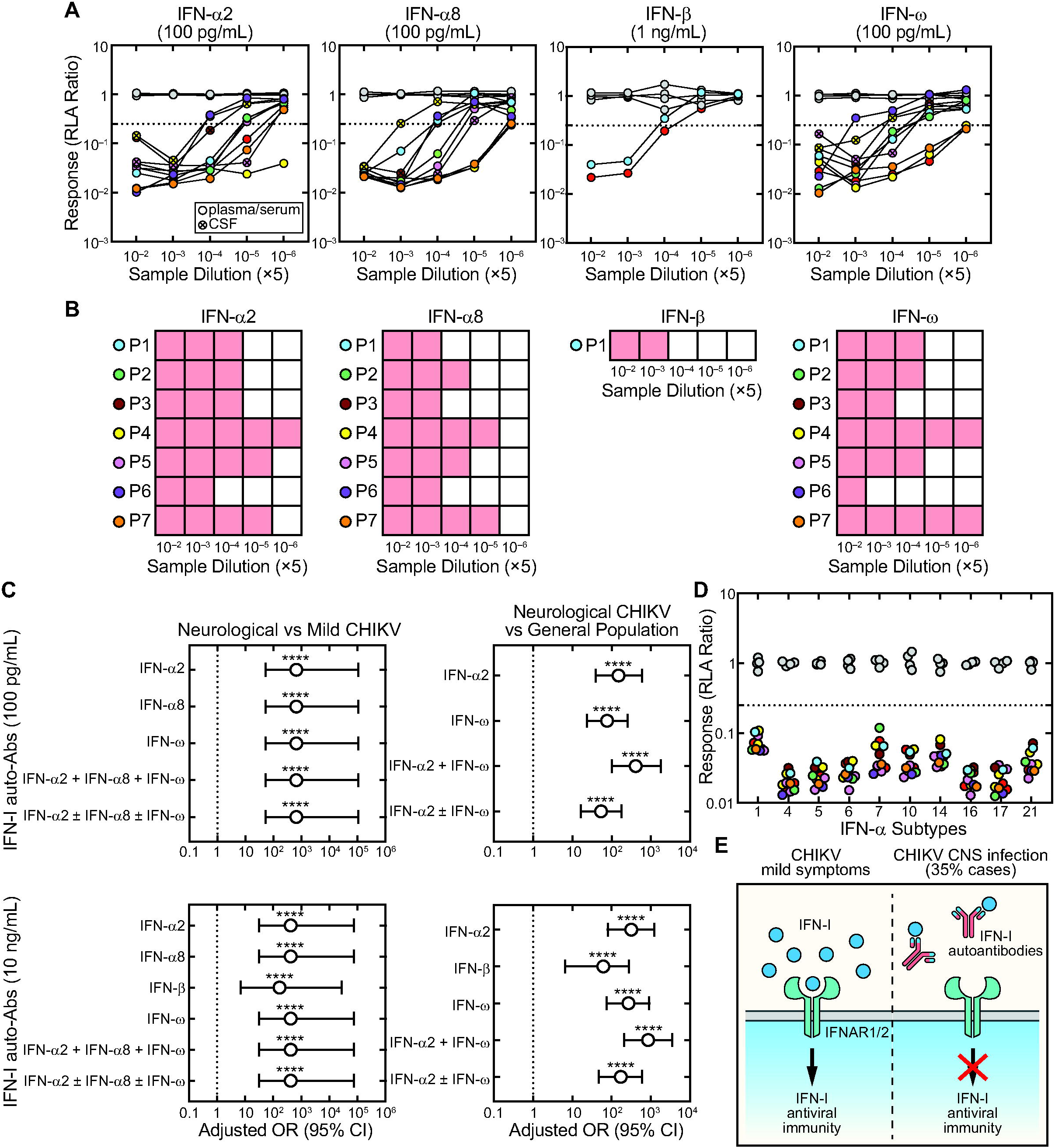
The potency and diversity of IFN-I autoantibodies in patients with severe CHIKV-infection. (*A*) Luciferase-based neutralization assay on serially diluted patient biofluids in the presence of constant amounts of IFN-I; RLA: relative luciferase activity. Samples with an RLA ratio < 0.25 were considered neutralizing. Gray: healthy donors; Red: IFN-I auto-Ab^+^ controls. (*B*) Heatmap depicting the neutralization activity of diluted samples containing IFN-I auto-Abs; red: positive, white: negative. (*C*) Odds ratios (OR) for the presence of IFN-I auto-Abs in CHIKV-infected patients with neurological complications relative to patients with milder CHIKV-infection (left) or the general population (right), with adjustment for sex and age. Open circle represents the OR and vertical bars represent the upper and lower limits of the 95% confidence interval. \*\*\*\**P* < 0.0001. (*D*) Luciferase-based neutralization assay for auto-Abs against 10 IFN-α subtypes (100 pg/mL). Gray: healthy donors; Red: IFN-I auto-Ab^+^ control. (*E*) Model of auto-Abs neutralizing IFN-I in a subset of CHIKV-infected individuals causing defective antiviral IFN-I immunity and severe clinical outcomes. CHIKV-infected individuals without IFN-I auto-Abs retain normal IFN-I-mediated antiviral immunity.

### Risk of CHIKV Disease in Individuals with IFN-I Autoantibodies

To determine the risk of CHIKV diseases by carrying blood AAN-I-IFN, we estimated the odds ratios (OR) by logistic regression with adjustment for sex and age. Circulating AAN-I-IFN were found exclusively in patients with severe acute CNS CHIKV infection (*P* < 0.0001, Fisher’s exact test), with a risk of severe disease ∼400 (OR = 424.4, 95% CI = 32.1−74438.1, *P* = 8.2×10^−8^ for 10 ng/mL IFN-I) to ∼600 (OR = 653.3, 95% CI = 52.2−106512.8, *P* = 1.6×10^−9^ for 100 pg/mL IFN-I) times higher than in patients with milder CHIKV infection (Fig. 2*C*). Previously, we demonstrated that the IFN-α2 and IFN-ω auto-Abs occurs in 0.17% and 1.4% of healthy individuals under and over 70 years of age, respectively, in the general population (25). In this study, neither healthy individuals nor 225 CHIKV-infected patients with mild symptoms carried IFN-I auto-Abs. By using the published dataset (25) as the reference population for comparison, we found that the risk of CHIKV-related neurological complications was about 60 times higher (OR = 61.8, 95% CI = 6.5−278.0, *P* = 0.0025) in individuals carrying IFN-β auto-Abs, ∼300 times higher (OR = 317.6, 95% CI = 80−1218.0, *P* = 2.2×10^−11^) in those with IFN-α2 auto-Abs, ∼270 times higher (OR = 269.9, 95% CI = 75.6−914.0, *P* = 2.0×10^−11^) in those with IFN-ω auto-Abs, and ∼850 times higher (OR = 854.6, 95% CI = 209.9−3491.9, *P* = 1.1×10^−13^) in the presence of both IFN-α2 and IFN-ω auto-Abs capable of neutralizing 10 ng/mL IFN-I, than in the general population (Fig. 2*C*). The reference population is predominantly of European descent, whereas patients in the current study were from Martinique (Caribbean) and Brazil. Population-level variation in IFN-I autoantibody prevalence across ethnicities has been reported (26, 27), and the true background rate in Caribbean and Brazilian populations is unknown. This difference in ancestry between the current cohort and the reference population is acknowledged as a limitation of the comparison.

### Autoantibodies Neutralizing the 12 IFN-**α** Subtypes

IFN-α2 and IFN-α8 belong to the IFN-α family, which contains 12 subtypes, all of which use the same IFNAR1/2 receptor complex for signaling but with different binding affinities and biological potencies (28–31). The biofluids of eight patients were able to neutralize both IFN-α2 and IFN-α8. We therefore tested whether other IFN-α subtypes were also neutralized to a similar extent. We preincubated patient biofluids positive for IFN-α2 and IFN-α8 auto-Abs, with 100 pg/mL IFN-α1, IFN-α4, IFN-α5, IFN-α6, IFN-α7, IFN-α10, IFN-α14, IFN-α16, IFN-α17, or IFN-α21 for one hour before AIR cell treatment for 24 h. All samples effectively neutralized all other members of the IFN-α family (Fig. 2*D*). Interestingly, the neutralization activity of CSF samples from P4 and P5 was similar to that of the corresponding serum for all IFN-α subtypes, consistent with in the data previously obtained for diluted samples. These data are consistent with the results of previous studies in which IFN-α auto-Abs, once developed, were shown to neutralize other subtypes of IFNα (14–16, 32).

## Discussion

In conclusion, we report that AAN-I-IFN underlie severe CNS CHIKV infection in ∼35% of cases studied. These auto-Abs, with a high neutralization capacity, circulate in both blood and CSF. They effectively neutralize high concentrations (from 1 to 1000 ng/mL) of IFN-α2, IFN-α8, and IFN-ω, and all IFN-α subtypes in samples diluted 1:20. The auto-Abs were detected in both serum and CSF samples from both patients for which both samples were available, consistent with our previous studies reporting the detection of IFN-I auto-Abs in the CSF of most West Nile virus encephalitis patients with AAN-I-IFN in the blood. We previously showed that AAN-I-IFN are present before severe viral infection and are the cause rather than the consequence of severe viral infection (15, 16, 33). Together with the detection of AAN-I-IFN in patients with encephalitis following live attenuated CHIKV vaccination (20, 34), our findings support a model in which individuals with such auto-Abs are unable to mount a sufficient IFN-I response upon CHIKV infection, thereby rendering them highly susceptible to severe CNS CHIKV disease (Fig. 2*E*). The defective antiviral IFN-I immunity of these individuals may lead to excessive viral replication in the periphery and in target organs, notably resulting in severe CNS infection once CHIKV crosses the blood-brain barrier. Other cases of severe CHIKV CNS infection in individuals without AAN-I-IFN may be due to genetic defects impairing the IFN-I response or production pathways (17, 18).

These findings have important clinical and public health implications. Screening for AAN-I-IFN is warranted in areas in which CHIKV is endemic, particularly for individuals over the age of 65 years. P1 in this study was previously diagnosed with SLE, consistent with the finding of AAN-I-IFN in other SLE patients (35), suggesting that screening for AAN-I-IFN in people with known autoimmune conditions might be beneficial. Since P6 and P7 were relatively young and harbored highly potent AAN-I-IFN, it is tempting to speculate that they might have undiagnosed autoimmune diseases. Testing for AAN-I-IFN could be proposed before vaccination with live-attenuated vaccines, including CHIKV VLA1553 (20) and Yellow Fever Y17D (36). Such screening program would help identify carriers of AAN-I-IFN in the general population, and AAN-I-IFN carriers should strive to reduce their exposure to ticks and mosquitoes, especially in areas in which arboviruses, such as CHIKV, are endemic. Given the rarity of IFN-β auto-Abs, IFN-β treatment may be helpful for treating or preventing severe CHIKV disease, provided that it is administered early. Other therapeutic strategies potentially appropriate for use in AAN-I-IFN carriers include Chimeric Autoantibody Receptor T-cell therapy and decoy IFN-I (37, 38).

## Materials and Methods

### Patients

We enrolled four independent cohorts of 245 individuals infected with CHIKV from Martinique, France, (CARBO cohort, Clinicaltrials.gov: NCT01099852) and Brazil, and 10 healthy individuals from Brazil. The cases were aged 0 to 89 years; 62% were female and 38% were male. Written informed consent was obtained from each individual in accordance with the local regulations, with institutional review board (IRB) approval. CHIKV infection was diagnosed based on the presence of CHIKV-specific IgM antibodies and/or CHIKV RNA in the plasma, serum, or cerebrospinal fluid. Patient stratification was based on the severity of infection and clinical manifestations, including mild symptoms, encephalitis, myelitis, and encephalomyelitis. The experiments were conducted in France and the USA in accordance with local regulations and guidance from the IRB committees of the Imagine Institute in Paris, France, and The Rockefeller University in New York, USA.

### IFN-I neutralization assay

We assessed the levels of autoantibodies against IFN-I by neutralization assays, as previously described (23). In brief, 50,000 AIR cells per 100 μL Dulbecco’s Modified Eagle’s Medium (Gibco, 10566016) containing 10% fetal bovine serum and 1% penicillin/streptomycin were dispensed into the wells of a 96-well plate and incubated for 24 h. Patient plasma, serum, or CSF (1:20 dilution) samples were incubated with 100 pg/mL IFN-α2, IFN-α8, or IFN-ω, or 1 ng/mL IFN-β in the presence of EnduRen substrate (Promega, E6482) for one hour with shaking (600 rpm) at room temperature. Cells were washed with 100 μL phosphate-buffered saline and treated with the sample mixture for 24 h at 37°C in an incubator with an atmosphere containing 5% CO_2_. *Renilla* luciferase activity was quantified by measuring the luminescence signal and normalizing against the median of all the samples tested. The 12 IFN-α subtypes were obtained from PBL Assay Science (11002). Samples with a relative luciferase activity ratio < 0.25 were considered to be neutralizing based on the results for a large cohort of healthy donors in previous studies (20, 23).

### Statistical analysis

Fisher’s exact test was performed to compare IFN-I auto-Ab levels between the encephalitis/myelitis and the mild infection group. The odds ratios and *P* values for the presence of IFN-I auto-Abs (neutralizing 100 pg/mL for IFN-α2, IFN-α8, and/or IFN-ω or 10 ng/mL for IFN-α2, IFN-α8, IFN-β, and/or IFN-ω) in patients with CHIKV-related neurological complications relative to those with mild CHIKV infection or the general population were estimated by Firth bias-corrected logistic regression, as implemented in the logistf R package, with adjustment for sex and age. Patients for whom plasma or serum samples were tested were included in the analysis. Statistical significance is indicated in the figure legend, as appropriate. **** for *P* < 0.0001.

## Data Availability

All data produced in the present study are available upon reasonable request to the authors.

## Acknowledgments

We thank the patients and their families for participating in our research. We thank all members of both branches of the Laboratory of Human Genetics of Infectious Diseases for discussions and technical and administrative support. We thank Lazaro Lorenzo and Yelena Nemirovskaya for administrative assistance. Funding: The Laboratory of Human Genetics of Infectious Diseases is supported by the Howard Hughes Medical Institute, The Rockefeller University, the St. Giles Foundation, the Stavros Niarchos Foundation (SNF) as part of its grant to the SNF Institute for Global Infectious Disease Research at The Rockefeller University, the National Institutes of Health (NIH) (R01AI163029), the National Center for Advancing Translational Sciences (NCATS), the NIH Clinical and Translational Science Award (CTSA) program (UL1TR001866), the French *Agence Nationale de la Recherche* (ANR) under the France 2030 program (ANR-10-IAHU-01), the HORIZON-HLTH-2024-DISEASE-08-20 program under GA 101191725 (InFlaMe), the ANRS projects DéméléJEV (ANRS0629) and LSDengue (ANRS-23-PEPR-MIE-0007), the Integrative Biology of Emerging Infectious Diseases Laboratory of Excellence (ANR-10-LABX-62-IBEID), the French Foundation for Medical Research (FRM) (EQU202503020018), ANR GENVIR (ANR-20-CE93-003), ANR AI2D (ANR-22-CE15-0046), the HORIZON-HLTH-2021-DISEASE-04 program under grant agreement 101057100 (UNDINE), the ANR-RHU COVIFERON Program (ANR-21-RHUS-0008), the Square Foundation, *Grandir – Fonds de solidarité pour l’enfance*, the *Fondation du Souffle*, the SCOR Corporate Foundation for Science, the Battersea & Bowery Advisory Group, William E. Ford, General Atlantic’s Chairman and Chief Executive Officer, Gabriel Caillaux, General Atlantic’s Co-President, Managing Director and Head of Business in EMEA, and the General Atlantic Foundation, the French Ministry of Higher Education, Research, and Innovation (MESRI-COVID-19), *Institut National de la Santé et de la Recherche Médicale* (INSERM), REACTing-INSERM, Paris Cité University, and the Imagine Institute. P.B. was supported by a “Poste CCA-INSERM-Bettencourt” (with support from the Bettencourt-Schueller Foundation), and the FRM (EA20170638020). A.G. was supported by a French National Agency for Research grant as part of the “Investissement d’Avenir” program (ANR-10-LABX-62-01). The authors thank the Centre de Ressources Biologiques de Martinique (CeRBiM) CHU Martinique, France, ID number BRIF BB-0033-00099, for managing patient samples. The CARBO cohort was supported by a grant from the Clinical Research Hospital Program from the French Ministry of Health (PHRC “2009”).

## Author Contributions

V.L.T., A.G., M.M.C., A.C., S.Y.Z., and J.L.C. designed research; V.L.T., A.G., M.M.C., M.P.S.S., S.B., I.C., M.S.R., L.S., J.D.S., P.A.A., A.P., P.B., L.A., C.P., R.F.O.F., I.C.S., A.C., and A.C. performed research; V.L.T., A.G., M.M.C., A.C., S.Y.Z., and J.L.C. analyzed data; and V.L.T., A.G., A.C., S.Y.Z., and J.L.C. wrote the paper.

## Classification

Biological Sciences, Medical Sciences

